# Tinnitus: An Unrecognised Symptom of Functional Neurological Disorder

**DOI:** 10.64898/2026.03.16.26348516

**Authors:** David D.G. Palmer, Mark J. Edwards, Jason B. Mattingley

## Abstract

**Background:** Functional neurological disorder (FND) is a common neurological condition characterised by symptoms which vary characteristically with attention. In the sensory realm, these symptoms frequently take the form of ‘phantom’ perception in the absence of sensation. While the condition is generally regarded not to cause auditory symptoms, tinnitus is a phantom perception which varies with symptom-focused attention, and is suggested to have similar underlying mechanisms to those proposed for FND. Based on this, we hypothesized that tinnitus might reflect the same underlying process as FND, and that it would therefore be more common in people with FND (pwFND).

**Methods:** Using an international database, we compared the proportions of pwFND who reported tinnitus with a control group. To ensure that observed differences were not attributable to agreement bias in symptom reporting, we also conducted an experiment where pwFND and controls were asked to report which symptoms they had experienced in the past month, 14 of which were symptoms of FND, and 7 of which were unrelated.

**Results:** Rates of tinnitus were significantly higher in the FND group (54% HDI 50 – 57%, n=732) than the control group (17% HDI 8.5 – 25%, n=59). In the symptom reporting experiment, pwFND (n=38) reported more FND-related symptoms than controls (n=38), but there was no between-group difference in reporting of non-FND related symptoms.

**Discussion:** Based on the markedly higher prevalence of tinnitus in pwFND than controls, and the substantial overlap in mechanisms and phenomenology, we believe tinnitus should be considered a possible symptom of FND, where both conditions reflect a failure of symptom resolution after incitement by a peripheral stimulus.

## Introduction

Functional neurological disorder (FND) is one of the most common causes of symptoms seen in clinical neurology.^1,2^ Symptoms of the condition are protean, and can encompass almost any class of symptom seen in other neurological diseases. They are distinguished by their striking modulation by attention, worsening with symptom-focused attention, and improving, or even temporarily ceasing, when attention is directed elsewhere. Often, FND begins in the setting of another illness,^3^ and in this setting the presenting symptoms may be similar to the symptoms of the inciting condition.

Current understanding of functional neurological symptoms conceptualises them as abnormal outcomes of otherwise normal brain processes.^4^ This account is most often framed within the Bayesian brain model, in which perception and action arise from hierarchical inference. At each level of this hierarchy, expectations (priors) are compared with incoming sensory information (likelihoods), and beliefs are updated according to their relative precision. When prior expectations are assigned high confidence, they may persist despite contradictory sensory evidence; conversely, when priors are uncertain, they may be rapidly revised in response to reliable incoming information. Importantly, the hierarchical organisation of inference means that expectations at one level provide the effective sensory input to the level above.

Within this framework, functional neurological symptoms are understood as arising from abnormal priors of perception or function that fail to update appropriately in response to sensory evidence from the body.^4^ Crucially, such priors are not assumed to arise spontaneously, but rather to develop in contexts that signal potential dysfunction of a given faculty. This process is commonly observed in neurological practice in the phenomenon of so-called “functional overlay,” in which functional symptoms emerge in individuals with non-functional neurological conditions and closely mimic the underlying condition. These functional symptoms occur commonly in all people,^5^ but in those susceptible to FND the symptoms may not resolve when the inciting non-functional pathology improves, suggesting that FND reflects a failure of symptom resolution rather than abnormal symptom generation.

Tinnitus, “the conscious awareness of a tonal or composite noise for which there is no identifiable corresponding external acoustic source,”^6^ is a common “phantom” sensation. Contemporary models of tinnitus propose mechanisms that closely parallel those invoked in functional neurological symptoms, emphasising abnormalities in perceptual inference and the updating of sensory expectations.^7^ Like functional symptoms, tinnitus is strongly modulated by attention^8^ and, like functional sensory symptoms, is frequently triggered by related peripheral insults to the auditory system, and constitutes a positive perceptual experience in the absence of sensation. Finally, the conditions share Despite these theoretical and phenomenological parallels, conventional descriptions of FND do not include symptoms in the auditory domain.^9–11^ If auditory perception were genuinely spared in FND, this would imply that the pathophysiological processes underlying the disorder are not domain-general, and that important insights might be gained by contrasting affected and unaffected sensory systems.

However, given the overlap in proposed mechanisms between tinnitus and functional sensory symptoms—and the close resemblance in their phenomenology—we hypothesised that tinnitus may be more prevalent in people with FND (pwFND) than in the general population. If so, this would support the view that tinnitus falls within the symptom spectrum of FND and, more broadly, that FND reflects a domain-general disorder of perceptual inference characterised by impaired symptom resolution.

## Methods

Our study used data from FND Research Connect (www.fnd-research.org), an international database through which both people with FND (pwFND) and people who do not have FND (controls) can enrol and provide data for both anonymised use in research, and for recruitment as research participants in other studies.

Participants in the database update their information every four months, meaning that for most individuals, longitudinal data are available.

The FND Research Connect database asks participants to report the experience of tinnitus as part of a matrix of symptoms with the overall question, “Please tell us which symptoms of FND you have experienced”. The wording of the entry for tinnitus in this matrix is, “Tinnitus (ringing or buzzing in the ears)”. Participants endorsing the symptom can respond either “I have had this symptom in the last month” or “I have had this symptom in the past (more than two months ago)”. Participants who report having had tinnitus at any time are subsequently asked, “Do you have hearing loss due to past exposure to loud noise, ageing, or a condition affecting your ears?”. In analysing the data, we used only the data for symptom experience in the past month.

Data were analysed using Python, with Bayesian regressions performed using the Bambi package.^12^ We included all records from FND Research Connect with at least one complete response set for experienced symptoms. The proportions reporting tinnitus were compared using Bayesian logistic regression. Posterior means with 95% highest density intervals (HDIs) are reported, as well as interpretation with respect to a region of posterior equivalence (ROPE) defined as +/-0.1 standardised mean differences. Odds were calculated for each of the three portions of the posterior distribution (above the ROPE, within the ROPE, and below the ROPE) against its complement, and the largest of these posterior odds is reported.

To determine whether an apparent difference in prevalence of tinnitus might be due to a greater tendency to report symptoms by the FND group (i.e. an agreement bias), we subsequently performed an experiment nested within data collection for another project. As part of baseline data collection for the project, participants were asked to say whether they had experienced any of 14 symptoms of FND within the last month. To this, we added 7 further symptoms which have no overlap with FND (e.g. sore throat, swollen ankles, cough). We then performed a Bayesian Poisson regression comparing the number of symptoms from the FND-related and non-FND related groups reported by participants in the control group and the patient group. Bayesian hypothesis testing was performed using the same methodology described above, with the ROPE set as a difference of +/-1 reported symptom.

Anonymised data and analysis scripts are available through the Open Science Framework at https://doi.org/10.17605/OSF.IO/9CXJT.

## Results

We accessed the data of 791 registered participants from FND Research Connect, of whom 732 were pwFND and 59 were controls. Sixteen participants were excluded from the FND group because they were in remission from FND, making group allocation ambiguous. A total of 716 records for pwFND, and 59 for controls, were included in the analysis. Demographics of respondents are presented in Table 1.

**Table 1:**
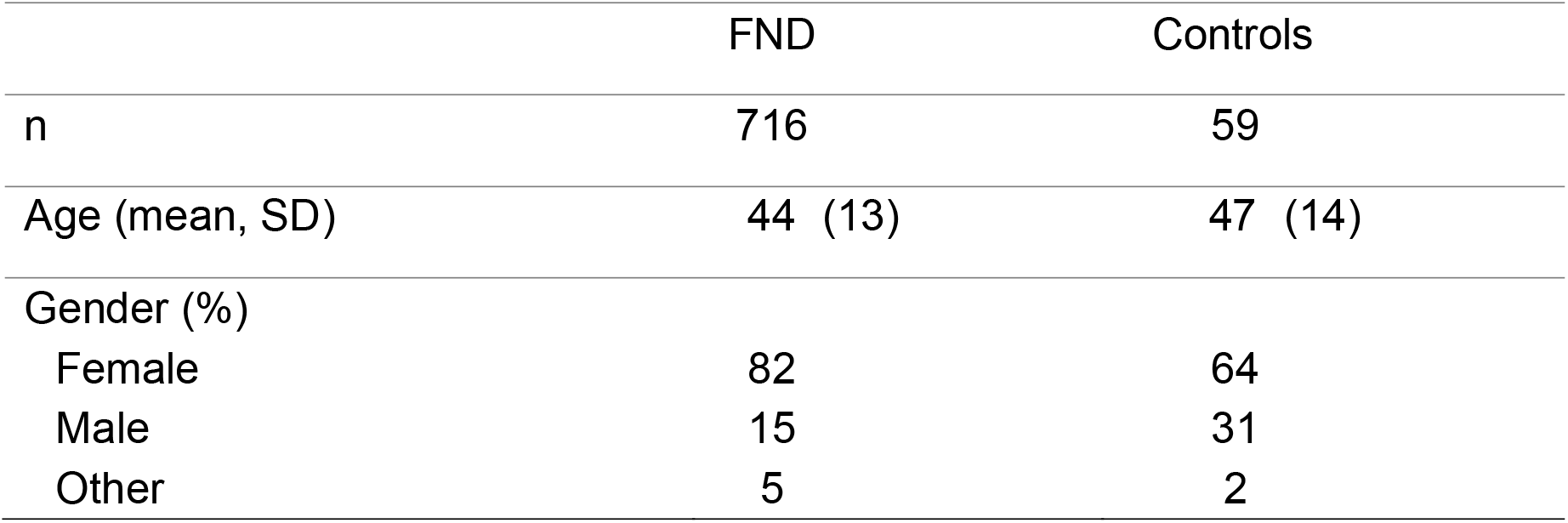
Demographics of participants included in analysis. Figures are rounded to 0 decimal places.

Results for the proportion of each group reporting tinnitus in the preceding month are presented in Figure 1A. 54% (HDI 50 – 57%) of the FND group and 17% (HDI 8.5 – 25%) of the control group reported experiencing tinnitus in the preceding month. The posterior odds for a between-groups difference were >4000. Of the participants who reported tinnitus, 27% (HDI 22 – 31%) of the FND group and 50% (HDI 25 – 76%) of the control group reported having hearing loss. The posterior odds for between-group difference in rate of hearing loss among those with tinnitus was 17.2.

**Figure 1.**
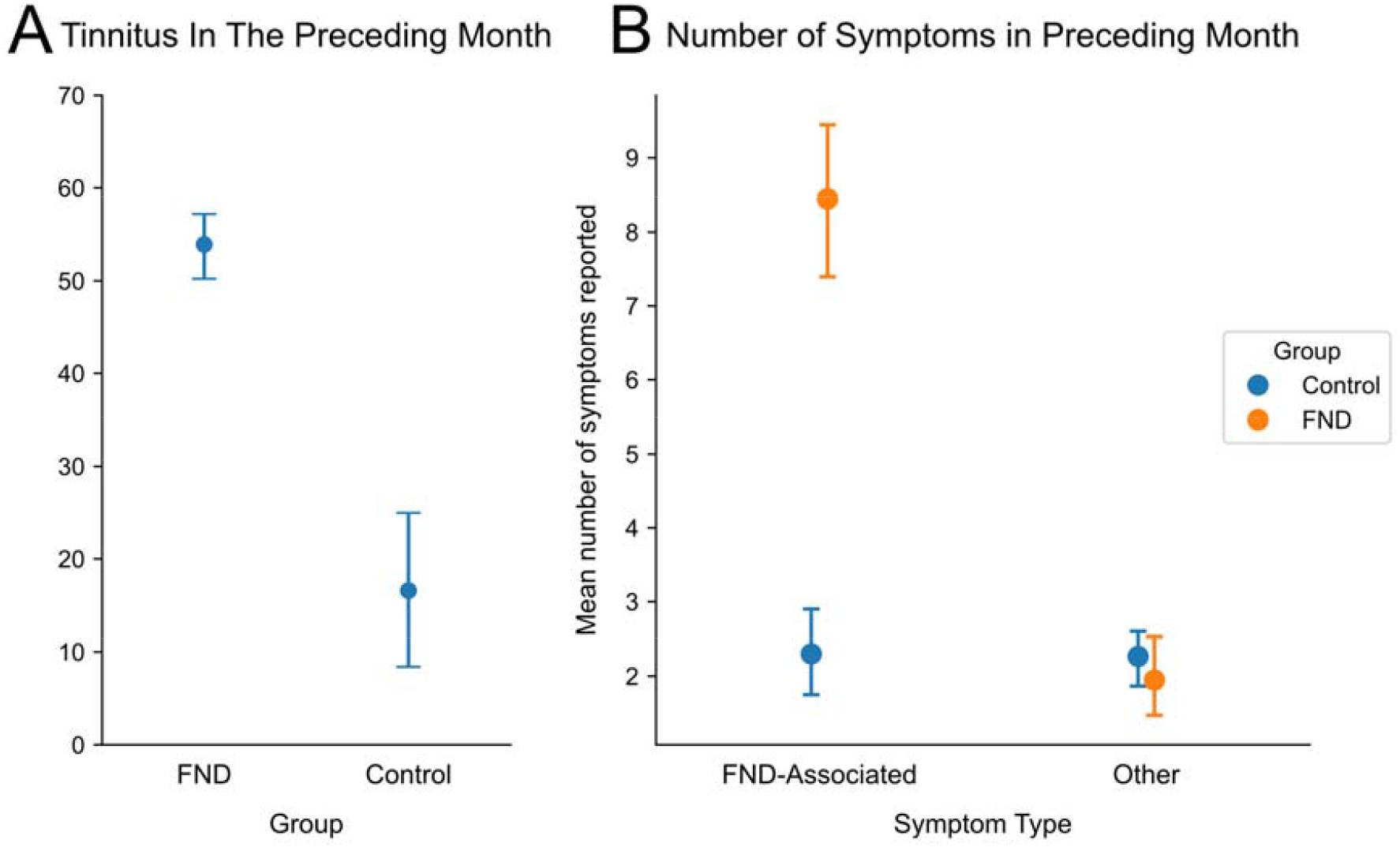
A. Posterior mean estimates with 95% HDI for proportion of each group who experienced tinnitus in the preceding month. B. Proportion of each group reporting FND-associated and non-FND associated symptoms in the preceding month.

38 pwFND and 38 controls participated in our experiment probing symptom-reporting. Figure 1B shows the number of FND-associated and non-FND associated symptoms reported by each group. As expected, the FND group reported substantially more FND associated symptoms in the preceding month than the control group, with posterior odds for between-group difference > 4000. By contrast, the mean number of non-FND associated symptoms reported by each group did not differ, with both groups reporting a mean of approximately 2 symptoms, with posterior odds for no between-group difference of 10.2.

## Discussion

We found that tinnitus was markedly more prevalent in people with functional neurological disorder (pwFND) than in controls drawn from the same database. More than half of pwFND reported experiencing tinnitus within the preceding month— approximately 6 times the rate at which controls experience the symptom (odds ratio: 5.7)—with overwhelming posterior evidence for a between-group difference. This frequency is comparable to the rates with which canonical symptoms of FND were reported. The difference is unlikely to be explained by a general tendency of pwFND to endorse symptoms, as a separate cohort of pwFND and controls showed strong evidence for no difference in rates of reporting of non-FND-related symptoms in an embedded symptom-reporting experiment. The rates of tinnitus in the control group are comparable with published estimates of prevalence in the general population.^13^

A key strength of this study is the use of a large, international cohort of pwFND with contemporaneous controls, collected using identical symptom wording and reporting methods. The inclusion of an internal test of agreement bias strengthens confidence that the observed association is specific to tinnitus rather than reflecting non-specific symptom endorsement. The Bayesian analytic framework allowed direct probabilistic statements about the magnitude and practical relevance of group differences.

Given their frequent co-occurrence and the significant overlap in their proposed mechanisms, we believe that tinnitus should be considered a symptom of FND. In addition to tinnitus being highly prevalent in pwFND relative to controls, the conditions share important phenomenological features with functional sensory symptoms: both are positive perceptual experiences occurring in the absence of corresponding peripheral input; both are strongly modulated by symptom-focused attention; and both are frequently triggered in the context of another relevant bodily insult. The convergence of research and theory on pathophysiological mechanisms for these two conditions also suggests shared pathophysiological mechanisms in abnormalities in perceptual inference and failure to appropriately update sensory expectations. The present results provide empirical support for linking these conditions at the clinical and theoretical levels.

Most episodes of tinnitus in the general population are transient, yet a subset of individuals develop persistent and distressing symptoms. This pattern accords with the view of FND as a disorder characterised not by symptom generation, but by failure of symptom resolution. On this account, both tinnitus and functional sensory symptoms may arise commonly following peripheral perturbations, but persist only when perceptual inference of dysfunction fails to normalise over time. The observation that pwFND who reported tinnitus in our study were significantly less likely to have hearing loss accords with this view. The tinnitus that these people experience could be conceived of as a failure of resolution of a symptom originally caused by a transient insult to the auditory system, while hearing loss might present an ongoing driver of tinnitus for those who have it.^14^

The strong association between tinnitus and FND also supports the broader view of FND as a domain-general disorder of inference rather than a syndrome confined to specific sensorimotor systems. While conventional descriptions of FND focus on motor, somatosensory, and visual symptoms, our findings suggest that auditory perception may be similarly affected. This raises the possibility that shared mechanisms—such as aberrant precision-weighting of sensory prediction error— may underlie symptoms across sensory domains.

Several limitations should be acknowledged. Our control group was substantially smaller than the pwFND group, reflecting the composition of the underlying database. The results for our control group were, however, consistent with published estimates of the prevalence of tinnitus, which increases our confidence in the result despite this limitation. The analysis was cross-sectional and based on symptom presence rather than onset, persistence, or severity, limiting causal inference. Additionally, participants in FND Research Connect may not be representative of all individuals with FND, particularly those with less severe or shorter-lived symptoms.

Future work should examine the longitudinal course of tinnitus in pwFND, including persistence, fluctuation, and resolution over time. Experimental paradigms probing auditory prediction-error processing and precision weighting could directly test proposed shared mechanisms. Finally, it would be informative to determine whether tinnitus in pwFND predicts broader sensory symptom burden, illness course, or response to treatment. Together, such work may help integrate tinnitus into a unified, mechanistic account of functional neurological disorder and inform cross-fertilisation of therapeutic approaches.

## Data Availability

Anonymised data and analysis scripts are available online through the Open Science Framework at https://doi.org/10.17605/OSF.IO/9CXJT

